# RARE CODING VARIANT ASSOCIATIONS WITH PRIMARY OPEN-ANGLE GLAUCOMA IN AFRICAN ANCESTRY:A MULTI-COHORT EXOME-WIDE META ANALYSIS

**DOI:** 10.64898/2026.02.25.26347141

**Authors:** Aude Ikuzwe Benigne Sindikubwabo, Yijun Fan, Yan Zhu, Lannawill Caruth, Rebecca Salowe, Bingxin Zhao, Joan O’Brien, Shefali S. Verma

## Abstract

Primary open-angle glaucoma (POAG) disproportionately affects individuals of African ancestry, yet rare coding variation in this population remains understudied. To address this gap, we performed a multi-cohort exome-wide meta-analysis across POAAGG, PMBB, All of Us, and UK Biobank, including 4,815 POAG cases and 22,922 controls of genetically inferred African ancestry. Although no gene reached exome-wide significance, we identified several suggestive gene-level associations driven by rare variants (minor allele frequency ≤0.1% or singletons),including signals in *SRF, BLTP3A, METTL2A*, and *KRT10*. Among these, *SRF* demonstrated the strongest association and was driven by rare missense variants with moderate effect sizes. Given its role in cytoskeletal organization and actin dynamics; processes central to trabecular meshwork function and intraocular pressure regulation *SRF* represents a biologically plausible candidate gene. Notably, these genes have not been previously highlighted in predominantly European ancestry POAG association studies, suggesting potential ancestry-specific rare variant contributions. Overall, our findings highlight the critical importance of investigating rare coding variation in POAG, in disproportionately affected populations to deepen understanding of POAG etiology and genetic risk.

## INTRODUCTION

Primary open-angle glaucoma (POAG) is an insidious neurodegenerative disease of the optic nerve (ON) that causes progressive vision loss.^1^ This disease affects 68.56 million individuals worldwide and is the leading cause of irreversible blindness in African ancestry individuals.^2, 3^Genetic factors are a major driver of POAG, with heritability estimates exceeding 70% in some populations.^4, 5^The majority of genetic research on POAG has focused on genome-wide arrays and imputation techniques, identifying hundreds of common variants with modest effect sizes.^6-10^ However, these techniques often fail to capture rare variants, which may contribute to a portion of the unexplained heritability of this disease.^11^ The identification rare, high-impact variants is essential to fully elucidate the genetic underpinnings of POAG and to improve risk prediction and therapeutic discovery, especially in underrepresented populations.

Approximately 3-5% of adult POAG is Mendelian and driven by highly penetrant mutations in genes such as *MYOC, OPTN*, and *TBK1*.^12^ Beyond these established genes, family-based and exome-wide burden sequencing studies have identified additional rare coding variants that contribute to the complex, polygenic form of POAG. Whole-exome sequencing (WES) in large, multi-generational families has revealed several causative genes associated with POAG.^13-17^ More recently, exome analysis in large biobanks, such as the UK Biobank, has confirmed known genes and uncovered novel candidates associated with POAG and intraocular pressure (IOP).^18-20^

The contribution of rare variants to POAG in African ancestry individuals remains poorly understood, despite their disproportionate burden of disease. In one study in African Americans and Ghanaians, exome array analysis identified several rare variants associated with POAG, including a variant in the *TMCO4* gene.^21^ Additionally, WES in an African American family identified a novel missense variant in the *EFEMP1* gene.^22^ However, large-scale sequencing studies in African ancestry populations remain limited. This ancestry group harbors the highest global levels of genetic diversity and may carry rare alleles that are rare or absent in non-African populations, highlighting the importance of their inclusion in such studies.^23, 24^

In this study, we aimed to identify rare variants associated with POAG in individuals of African ancestries. We first conducted WES on African ancestry individuals from the Primary Open-Angle African American Glaucoma Genetics (POAAGG) study and performed gene-based burden testing on exome sequencing data. We then meta-analyzed these findings with African ancestry individuals in several large biobanks, including Penn Medicine Biobank (PMBB), All of Us (AoU), and UK Biobank (UKBB).Our analyses identified suggestive rare variant associations in several genes, including *SRF, BLTP3A, METTL2A*, and *KRT10*, which have not been highlighted in prior rare variant studies focused on predominantly non-African ancestries. These findings emphasize the value of ancestry-specific exome analyses and support the contribution of rare coding variation to POAG risk in populations that are disproportionately affected by the disease.

## METHODS

### Study Population and phenotype definition

#### POAAGG

The POAAGG cohort consists of self-identified Black participants (African American, African descent, African Caribbean individuals), aged 35 years and older, who were recruited from the Philadelphia area between 2010 and 2019. Subjects were enrolled during routine ophthalmology visits at the University of Pennsylvania and other sites.^25, 26^ At enrollment, a glaucoma specialist or ophthalmologist classified each individual as a POAG case, suspect, or control according to established diagnostic criteria.^25^ Written informed consent was obtained from all participants. The study was approved by the University of Pennsylvania Institutional Review Board (protocol #812036) and was conducted in accordance with the principles of the Declaration of Helsinki.

#### PMBB

The Penn Medicine Biobank (PMBB) is a large, health system-based biorepository embedded within the University of Pennsylvania.^27^ To date, approximately 350,000 individuals have consented to participate, of whom 57,170 have genome-wide genotype data imputed to the TOPMed reference panel and linked to their clinical records. In addition to genomic data, the PMBB integrates comprehensive electronic health record (EHR) information, including diagnostic codes, laboratory values, imaging results, medication histories, and selected lifestyle factors. A major strength of the PMBB is its substantial ancestral diversity, with approximately 30% of participants identifying as non-European ancestry, of which 17% are identified as African American. This diversity enhances the representativeness of the cohort and improves the generalizability of genetic research findings across populations.^27^

#### All of Us

The *All of Us* Research Program (AOU) is a large, prospective, longitudinal cohort study led by the National Institutes of Health with the aim of enrolling at least one million participants across the United States to advance precision medicine and accelerate biomedical discovery. To date, more than 600,000 individuals have enrolled, with over 400,000 participants having short-read whole-genome sequencing data (https://databrowser.researchallofus.org/). A distinguishing feature of the AOU cohort is its emphasis on diversity: approximately 77% of participants come from communities historically underrepresented in biomedical research.^28^ In addition to genomic data, the program integrates electronic health record (EHR) data from approximately 50 participating healthcare organizations. At each site, clinical data are harmonized to the Observational Medical Outcomes Partnership (OMOP) Common Data Model and transferred quarterly to the All of Us Data and Research Center for integration, curation, and periodic release to the research community.

#### UKBB

The UK Biobank (UKBB) is a large-scale, population-based research resource that provides extensive data for studying a broad range of health and disease outcomes.^29^ It includes approximately 500,000 participants from across the United Kingdom, with linked EHR and comprehensive genome-wide genetic data. Genotyping was performed using the Applied Biosystems UK BiLEVE and UK Biobank Axiom Arrays, followed by centralized genotype calling and rigorous quality control procedures. These arrays directly assay were then imputed using the Haplotype Reference Consortium and the combined UK10K and 1000 Genomes reference panels expanded the dataset to more than 90 million variants for analysis.^29^

### PHENOTYPING

Across PMBB, AOU, and UKBB, analyses were restricted to genetically inferred African ancestry individuals aged ≥35 years. Ancestry was determined using principal component analysis (PCA), and related individuals were retained. POAG cases were defined using harmonized ICD-based criteria across cohorts. Specifically, cases included individuals with ICD-9 codes 365.10 or 365.11, or any ICD-10 H40.11 variant (primary open-angle glaucoma). Individuals with ICD-10 codes H40.01* (borderline findings) and H42* (glaucoma in diseases classified elsewhere) were excluded to remove secondary glaucoma due to trauma, inflammation, drug effects, or other underlying conditions. In AOU, initial case identification incorporated relevant SNOMED codes (glaucoma and primary open-angle glaucoma) prior to ICD filtering. In UKBB, cases were additionally identified through self-report, including selection of “glaucoma” in questionnaire fields (data field 6148 or 20002) or a reported history of glaucoma surgery or laser treatment.

Controls were defined consistently across cohorts as individuals with no evidence of glaucoma, including no ICD-9 codes beginning with 365 and no ICD-10 codes within the H40 or H42 ranges. Additionally, controls in all three cohorts were required to have a record of at least one ophthalmology visit or report no recorded eye problems, in order to reduce the frequency of undiagnosed glaucoma among controls. This was defined as follows: In PMBB, controls had at least one prior ophthalmology visit at the University of Pennsylvania. In AOU, controls were required to have no relevant SNOMED codes (including POAG, open-angle glaucoma, or glaucoma suspect) and to report at least one eye doctor visit within the past 12 months (“*What is the total number visits to an eye doctor that you made in the last 12 months?*”). In UKBB, controls responded “No” to the glaucoma questionnaire item (field 6148, “*Has a doctor told you that you have any of the following problems with your eyes?”*) and did not report any eye problems in the verbal interview (data field 20002). Similar definitions have been used in prior large-scale glaucoma genetic studies.^9, 18, 30-34^

### EXOME DATA PROCESSING AND QUALITY CONTROL

#### PMBB

Sample and variant quality control (QC) were implemented following established best practices. At the sample level, quality control included exclusion based on sex discordance, low genotyping call rate (>99% required), and high heterozygosity or contamination, as well as removal of duplicate samples. Genetically inferred ancestry was determined using principal component analysis (PCA) to confirm sample ancestry and adjust for population structure. For whole-exome sequencing, samples with sex errors, excessive heterozygosity/contamination (D-stat > 0.4), or low sequence coverage (<85% of targeted bases at ≥20×) were excluded. For variant QC, genotype data underwent filtering for variant and genotype call rate (>99%), imputation quality (R^2^ > 0.7), removal of palindromic and non-biallelic sites, Hardy–Weinberg equilibrium test (p > 1 × 10^−8^), and minor allele frequency thresholds for common variant analyses. For exome sequence variants, low‐depth genotypes (DP < 7 for SNVs, DP < 10 for INDELs) were set to missing, and allele balance filters were applied (SNV AB ≥ 0.15; INDEL AB ≥ 0.20) to retain variants with acceptable heterozygote balance or homozygous alternate genotypes. Minor allele count (MAC) thresholds were applied as appropriate in downstream analyses

#### POAAGG

Initial quality control for sequencing data was done by Regeneron. Variant and genotype quality control (QC) were implemented using a two-step filtering process. First, at the sample level, genotype calls at variant sites were filtered based on depth of coverage (DP), where low-depth genotypes were set to missing (‘No-call’ or ‘./.’). Specifically, genotypes with DP < 7 for single nucleotide polymorphisms (SNPs) and DP < 10 for insertions/deletions (INDELs) were excluded. Following sample-level genotype filtering, variant sites were filtered based on allele balance (AB). Variant sites were omitted from the dataset if all samples carrying the variation had heterozygous (0/1) genotype calls and simultaneously failed the minimum AB cutoff. To be retained in the dataset, SNP variant sites required at least one sample to carry an alternate AB ≥ 0.15, and INDEL variant sites required at least one sample to carry an alternate AB ≥ 0.20. Moreover, Hardy–Weinberg equilibrium test (p > 1 × 10^−^15), genotype missingness filter (< 10%), and MAF cutoff (MAF < 0.01) have been applied before the downstream analyses.

#### AOU

Sequencing data were jointly called across all samples to generate a unified call set, improving variant accuracy through cross-sample quality control. Single-sample QC excluded samples with contamination, swaps, or preparation errors (research contamination threshold 3%). Additional joint-call QC incorporated hard filters, population outlier detection, allele-specific filtering, and sensitivity/precision evaluation. Cross-center batch effects were assessed and were minimal in high-confidence genomic regions. Variant-level quality control in All of Us included allele-specific Variant Quality Score Recalibration (VQSR), hard filtering for low-quality sites, excess heterozygosity, and removal of variants lacking high-quality genotypes (GQ ≥20, DP ≥10, AB ≥0.2). Joint calling across all samples enabled cross-sample refinement of variant accuracy, and batch effects across sequencing centers were formally evaluated.^36^

#### UKBB

Whole-exome sequencing of UK Biobank participants was performed at the Regeneron Genetics Center through a precompetitive collaboration among multiple pharmaceutical partners. Genomic DNA was sequenced using paired-end 75-bp reads with the IDT xGen v1 capture kit on the NovaSeq 6000 platform. Sequencing data were converted from BCL to FASTQ format and demultiplexed using 10-base barcodes (bcl2fastq v2.19.0). Initial quality control conducted by Regeneron included assessments of sex concordance, contamination, duplicate samples, and concordance with microarray genotyping data.^37^

### ANNOTATION AND GENE-BASED GROUP FILES

Variant annotation was performed using the Ensembl Variant Effect Predictor with Ensembl/GENCODE gene models^38^. Variants were annotated for predicted functional consequences and supplemented with additional functional annotations using the LoFTEE, dbNSFP, CADD and SpliceAI plugins. All annotations were performed against the GRCh38 reference genome. The complete annotation workflow and parameter settings are available in our VEP pipeline repository.All analyses were restricted to coding regions across datasets. To classify each variant, we used the following guidelines; high-confidence predicted loss-of-function (pLoF) variants were defined as those annotated as high-confidence by LOFTEE (LOFTEE HC). Damaging missense/protein-altering variants comprised variants not classified as high-confidence pLoF that met at least one of the following criteria: (i) annotated as missense, start-loss, stop-loss, or in-frame indel with REVEL ≥ 0.773 or CADD ≥ 28.1; (ii) SpliceAI delta score (DS) ≥ 0.2, where DS was defined as the maximum of DS_AG, DS_AL, DS_DG, and DS_DL; or (iii) low-confidence loss-of-function variants (LOFTEE LC). Other missense/protein-altering variants included missense, start-loss, stop-loss, and in-frame indels not meeting the damaging criteria above. Synonymous variants with SpliceAI DS < 0.2 were included as a control set.

REVEL and CADD thresholds were selected based on previously published cut-offs [36413997]. For gene-based testing, variants were grouped by annotated functional consequence and aggregated within genes. For each gene–variant set combination, association analyses were performed using minor allele frequency (MAF) thresholds of ≤1%, <0.1%, and <0.001%. Variants not meeting these frequency thresholds were considered ultra-rare and were analyzed as singletons.

### META-ANALYSIS

To combine association evidence across cohorts, we performed a two-tailed Stouffer’s z-score meta-analysis on the exome-wide association results. For each test (defined by gene, annotation group, and maximum minor allele frequency for burden tests), p-values from individual cohorts were converted to one-sided p-values and transformed into z-scores weighted by the square root of the effective sample size, where effective sample size was calculated as 2/((1/N_cases) + (1/N_controls)). Cohort-level weighted z-scores were summed and divided by the square root of the total effective sample size to obtain an overall meta-analytic z-score, from which a two-tailed meta-analysis p-value was derived. Results were filtered to retain associations meeting a Bonferroni corrected p-value threshold of 2.62e-06 and present in more than one contributing cohort.

### STATISTICAL ANALYSES

We used REGENIE^35^ to conduct the exome-wide association tests. Age, sex, and top 10 genetic PCs were adjusted as covariates. We applied Firth logistic regression to derive the summary statistics for our binary phenotype as recommended in Mbatchou *et al*.^*35*^ REGENIE employs a two-step procedure for testing rare variant associations. In step 1, common variants in the genotyping array data were included after quality control (MAC > 100, genotype missing rate < 10%, Hardy-Weinberg equilibrium *P* > 1×10^-15^) to capture the polygenic effects. The predictors from step 1 were then used in step 2 for variant-level and gene-based tests based on rare variants. For both levels of tests, we followed the default setting of REGENIE that only variants/burden masks with MAC > 5 were tested.

Exome-wide association analyses in the AoU biobank were performed using SAIGE-GENE^39^, analogous to our REGENIE-based analyses. Age, sex, and the top six genetic principal components were included as covariates. SAIGE step 1 was used to fit the null generalized mixed model using curated phenotype data and a sparse genetic relationship matrix (GRM), accounting for relatedness and case–control imbalance, following sample- and variant-level quality control as performed by AoU. In SAIGE step 2, gene-based association testing was conducted using the predefined variant categories described above. The complete analysis pipeline, including workflow configuration and execution scripts, is available in our GitHub repository (see code availability).

## RESULTS

A total of 4,815 POAG cases and 22,922 controls of African ancestry were included across four cohorts (PMBB, All of Us, POAAGG, and UK Biobank) (Table 1). Case definitions were based on electronic health record (EHR) algorithms in PMBB, All of Us, and UK Biobank, and clinician-adjudicated diagnoses in POAAGG. No gene reached exome-wide significance in the cross-biobank meta-analysis after Bonferroni correction; however, multiple genes demonstrated suggestive associations (Table 2). The strongest signal was observed in SRF (p = 6.32 × 10^−6^), driven by rare missense variants (max MAF = 0.001) with a positive effect estimate (β = 1.690, SE = 0.382). Additional suggestive associations included CRIP2 (p = 1.33 × 10^−5^), BLTP3A (p = 1.66 × 10^−5^), METTL2A (p = 2.42 × 10^−5^), RPL5 (p = 2.54 × 10^−5^), and KRT10 (p = 2.58 × 10^−5^). Notably, KRT10 was driven by an ultra-rare singleton missense variant. Other suggestive genes included TBC1D10B, CSHL1, GUCA2B, and UNC93A, with signals arising from low-frequency (MAF ≤ 0.01) missense or synonymous variant masks. Meta-analysis results of the top gene (*SRF*) in Figure 2, all additional results are provided in the Supplementary Tables.

**Table 1.** Summary of biobank cohorts included in the African ancestry exome-wide association study of primary open-angle glaucoma.

| Biobank | Cases | Controls | Total (N) | Cohort Ancestry | Case Definition | Genes Tested |
| --- | --- | --- | --- | --- | --- | --- |
| PMBB | 468 | 2,638 | 3,106 | AFR | EHR algorithm | 17,990 |
| All of Us | 1,472 | 8,368 | 9,840 | AFR | EHR algorithm | 19,027 |
| POAAGG | 2,542 | 2,542 | 10,910 | AFR | Adjudicated | 17,887 |
| UK Biobank | 333 | 3,548 | 3,881 | AFR | EHR algorithm | 17,820 |

AFR, African ancestry; EHR, electronic health record; POAAGG, Primary Open-Angle African American Glaucoma Genetics study. Case definitions: EHR algorithm = diagnosis codes plus medication criteria; Adjudicated = clinician-reviewed medical records. Genes tested refers to the number of protein-coding genes passing quality filters in each cohort.

**Table 2.** Top gene-level suggestive association results from cross-biobank meta-analysis of primary open-angle glaucoma in individuals of African ancestry.

| Cohort | Gene | p-value (Stouffer) | $\beta$ (SE) | Max MAF | Annotation Group |
| --- | --- | --- | --- | --- | --- |
| AFR | SRF | $6.32 \times 10^{-6}$ | 1.690 (0.382) | 0.001 | Other missense |
| AFR | CRIP2 | $1.33 \times 10^{-5}$ | -0.823 (0.193) | 0.01 | Synonymous |
| AFR | BLTP3A | $1.66 \times 10^{-5}$ | 0.559 (0.265) | 0.0001 | Other missense |
| AFR | METTL2A | $2.42 \times 10^{-5}$ | 2.005 (0.490) | 0.001 | Synonymous |
| AFR | RPL5 | $2.54 \times 10^{-5}$ | 0.866 (0.249) | 0.01 | Synonymous |
| AFR | KRT10 | $2.58 \times 10^{-5}$ | 1.361 (0.331) | Singleton | Other missense |
| AFR | TBC1D10B | $3.29 \times 10^{-5}$ | 0.049 (0.012) | 0.001 | Synonymous |
| AFR | CSHL1 | $3.52 \times 10^{-5}$ | 0.672 (0.153) | 0.01 | Synonymous |
| AFR | GUCA2B | $3.81 \times 10^{-5}$ | -0.638 (0.180) | 0.01 | Other missense |
| AFR | UNC93A | $4.89 \times 10^{-5}$ | -0.446 (0.228) | 0.001 | Synonymous |

Suggestive meta-exwas results are sorted by Stouffer combined p-value. $\beta$ , effect size estimate; SE, standard error; Max MAF, maximum minor allele frequency threshold used in burden mask; Singleton = ultra-rare variant not included in the other threshold. Annotation groups as defined in the method section

**Figure 1.**
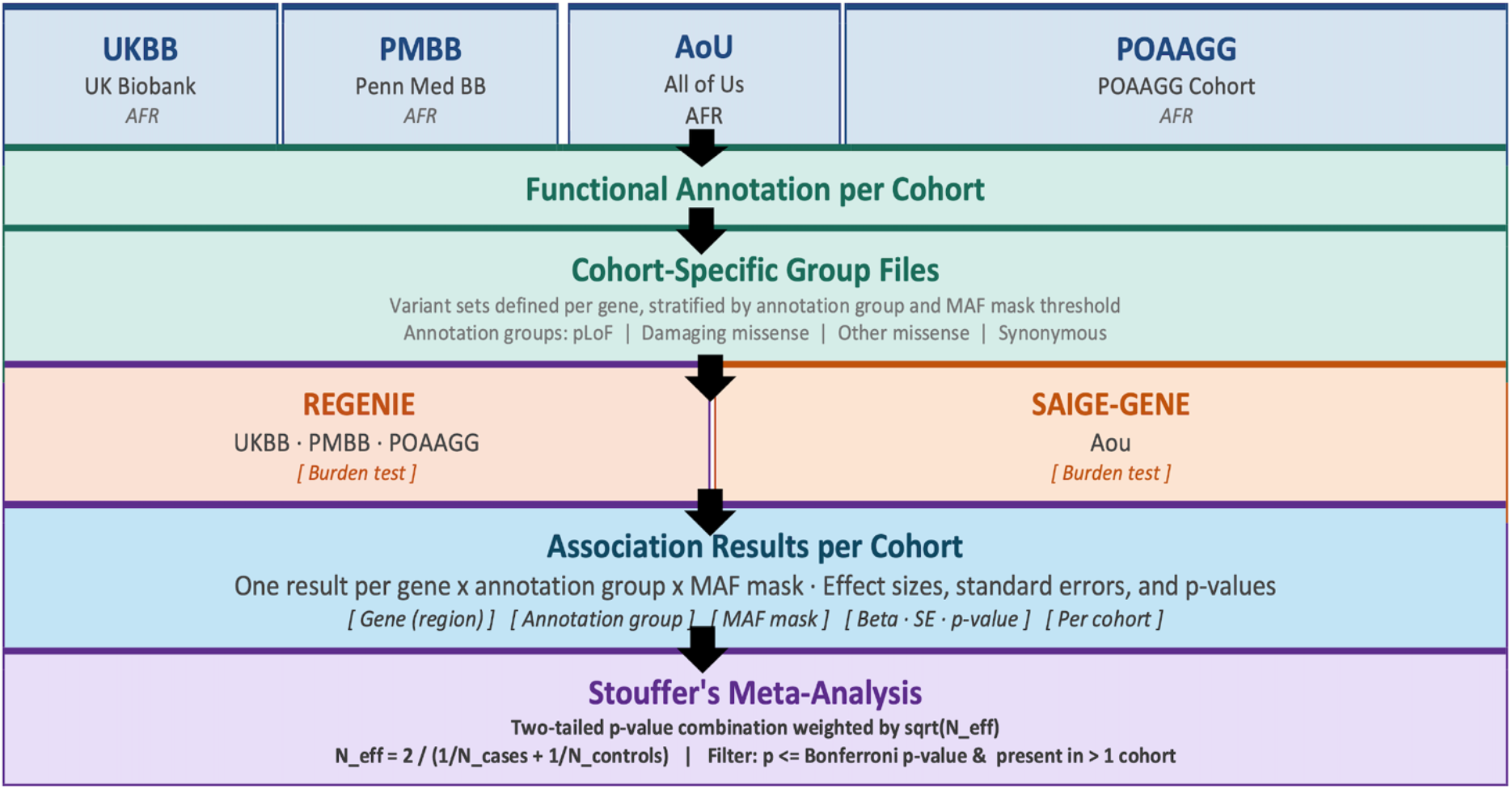
Overview of the ExWAS meta-analysis workflow.” Schematic illustrating the sequential steps of the exome-wide association study meta-analysis, from cohort-level variant filtering and single-variant/gene-based burden testing to cross-cohort meta-analysis.

**Figure 2.**
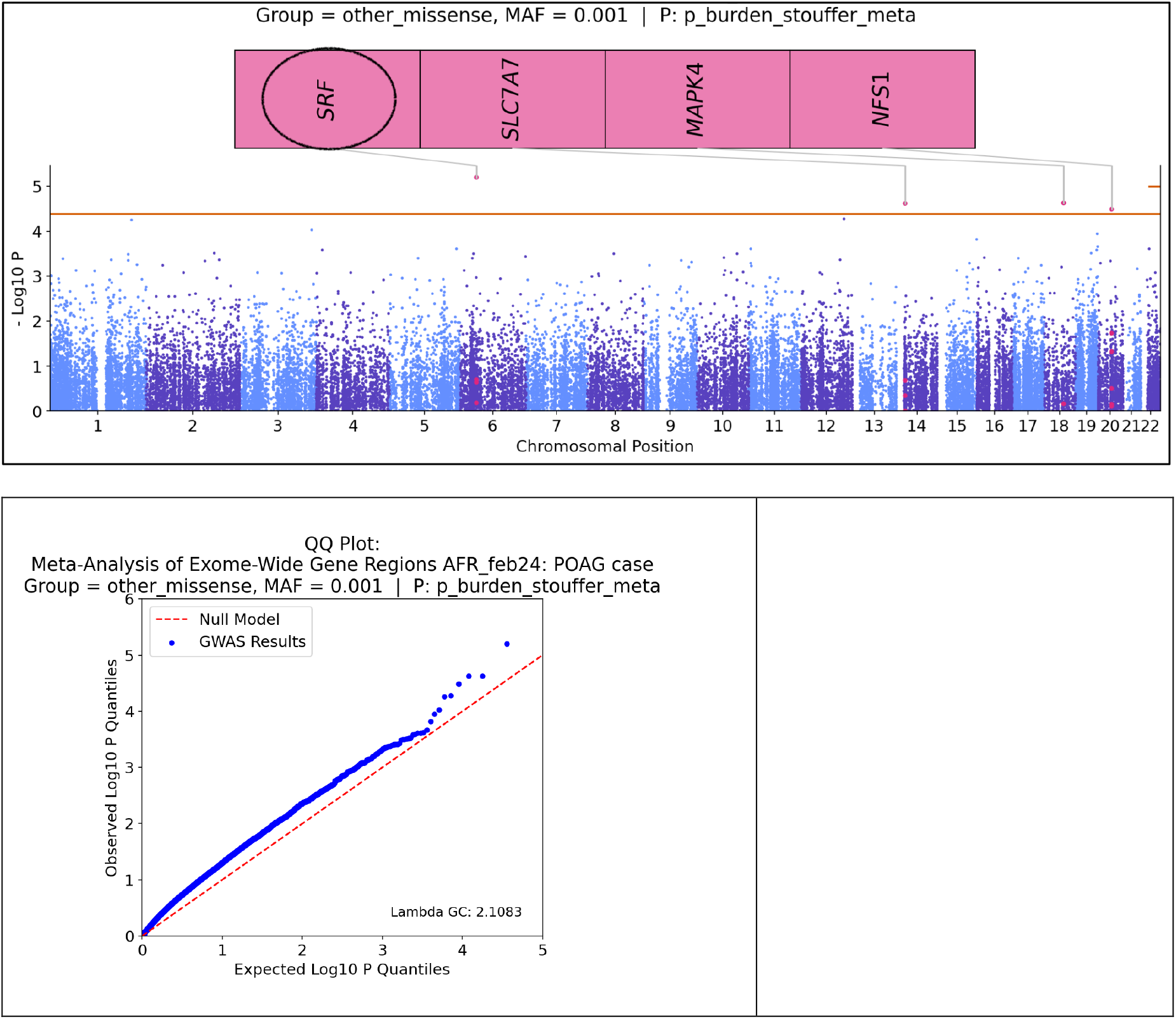
Manhattan and quantile-quantile (Q-Q) Manhattan and quantile-quantile (Q-Q) plots from the exome-wide association study (ExWAS) meta-analysis of primary open-angle glaucoma (POAG). The Manhattan plot displays –log_10_(P-values) for each gene-variant across the exome, with the horizontal dashed line indicating threshold (P < 0.0004). The Q-Q plot shows the observed versus expected – log_10_(P-values), with the genomic inflation factor (λ) indicating the degree of test statistic inflation

## DISCUSSION

In this African ancestry exome-wide meta-analysis of POAG, we identified several suggestive gene-level associations driven by rare coding variants, including signals in SRF, BLTP3A, METTL2A, and KRT10, among others. While no gene reached exome-wide significance, several top signals were based on rare variants (MAF ≤0.1% or singletons) and were observed across two to three cohorts, supporting their potential relevance. Notably, these genes have not been prominently implicated in prior predominantly European POAG GWAS, suggesting possible ancestry-specific rare variant contributions. Our findings therefore extend prior common-variant GWAS and exome studies by evaluating coding variation specifically within African ancestry populations, where rare allele spectra differ and may enable detection of signals not observed in other populations.

Among the top signals, SRF is particularly notable given its role in cytoskeletal organization and actin dynamics, processes central to trabecular meshwork function and aqueous humor outflow, implicating a potential mechanism related to intraocular pressure regulation. The association was driven by rare missense variants (MAF ∼0.1%) with relatively large effect size estimates, supporting functional relevance. BLTP3A, involved in lipid transport and membrane biology, may influence cellular stress or extracellular matrix processes relevant to optic nerve vulnerability. KRT10, driven by an ultra-rare singleton missense variant, encodes a structural keratin protein and may reflect epithelial or barrier-related mechanisms, although its role in ocular tissues requires clarification. Other genes such as METTL2A (RNA methyltransferase) and TBC1D10B (vesicle trafficking regulator) suggest potential involvement of transcriptional or intracellular signaling pathways not traditionally emphasized in POAG, pointing to possible novel mechanisms. Overall, the mixture of missense-driven signals and ultra-rare variants suggests heterogeneous functional effects that warrant targeted follow-up.

The top associations were observed across two to three cohorts, including both the deeply phenotype POAAGG clinic-based cohort and EHR-defined biobank cohorts (PMBB, AoU, UKBB). These differences in ascertainment, disease severity, and age distribution likely introduce heterogeneity in effect size estimates. Nevertheless, several genes, including *SRF and BLTP3A*, demonstrated contributions from more than one cohort, reducing the likelihood that signals are entirely cohort-specific artifacts. Some variability in effect magnitude was observed, which may reflect phenotype misclassification in biobank settings or limited power for ultra-rare variants. Ongoing sensitivity analyses, including leave-one-cohort-out meta-analyses, stricter case definitions, and exclusion of related individuals, will further clarify the robustness and stability of these findings across study designs.

By restricting analyses to genetically inferred African ancestry individuals across POAAGG, PMBB, AoU, and UKBB, we evaluated rare coding variation within a population that harbors greater genetic diversity and distinct rare allele spectra. Several top signals (e.g., SRF, BLTP3A, KRT10) were driven by low-frequency or ultra-rare variants (MAF ≤0.1% or singletons), variants that may be rare or absent in non-African datasets. Studying AFR cohorts therefore improves the ability to detect ancestry-specific rare coding contributions to POAG and addresses the underrepresentation of African ancestry individuals in prior sequencing-based glaucoma studies.

Our study follows the framework established by large-scale exome sequencing analyses of intraocular pressure and glaucoma, including Gao et al.^18^, who used gene-based rare variant aggregation in UK Biobank to identify coding variants influencing IOP with downstream relevance to POAG. Similar to that study, we applied burden-based testing across functional masks and focused on aggregated rare coding variation rather than relying solely on single-variant signals. In addition, consistent with large biobank exome sequencing studies demonstrating allelic series within genes (e.g., protein-altering variants in ANGPTL7 associated with lower IOP^20^), our top findings were gene-level signals driven by multiple rare coding variants with moderate effect sizes. While prior work has largely focused on predominantly European ancestry populations, our analysis extends this established rare-variant discovery paradigm to African ancestry POAG, emphasizing the importance of ancestry-specific exome evaluation.

Several limitations should be considered. First, phenotype definitions differed across cohorts, with POAAGG using adjudicated clinical diagnoses and biobank cohorts relying primarily on EHR- and self-report–based algorithms, which may introduce misclassification and attenuate effect estimates. Second, despite meta-analysis across four cohorts, power remains limited for ultra-rare variants, and no gene reached exome-wide significance; results may be sensitive to variant masking strategies and minor allele count thresholds. Third, differences in exome capture platforms, sequencing pipelines, and variant calling across cohorts may introduce residual batch effects despite quality control harmonization. Finally, our AFR-only design was intentional to prioritize ancestry-specific discovery, but replication in independent African ancestry datasets remains limited; future studies incorporating additional AFR cohorts and cross-ancestry analyses will be important to confirm and generalize these findings.

Future efforts should prioritize replication of these candidate genes in independent African ancestry glaucoma cohorts and consortia, including GGLAD and ADAGES, to validate ancestry-specific rare variant signals. Functional follow-up will be essential to clarify biological mechanisms, including evaluation of gene expression in ocular tissues, colocalization with regulatory signals, and experimental assessment of predicted loss-of-function and damaging missense variants using cellular or animal models. In the longer term, incorporation of rare variant burden into integrated risk frameworks, combining polygenic risk scores with rare coding variation and imaging or clinical biomarkers, may enhance disease risk stratification in African ancestry populations, pending robust replication and functional validation.

In summary, this multi-cohort whole-exome sequencing meta-analysis represents one of the largest rare coding variant studies of POAG conducted exclusively in African ancestry individuals. Although no gene reached exome-wide significance, we identified multiple biologically plausible candidate genes supported by aggregated rare coding variation across cohorts. These findings expand the investigation of POAG genetic architecture beyond common-variant GWAS and underscore the importance of ancestry-specific sequencing efforts. Continued replication and functional validation will be essential to clarify the contribution of rare coding variants to POAG risk in African ancestry populations.

## Data Availability

All data produced in the present study are available upon reasonable request to the authors

## Data and Code Availability

The SAIGE-GENE analysis pipeline used for the All of Us exome-wide analyses is publicly available at: https://github.com/PMBB-Informatics-and-Genomics/pmbb-nf-toolkit-saige-family/tree/main

## Supplementary information

Supplementary information accompanies this paper. All supplementary figures and tables are available at: https://github.com/Setia-Verma-Lab/POAG_EXWAS_META_SUPP

